# Isolation thresholds for curbing SARS-CoV-2 resurgence

**DOI:** 10.1101/2020.11.20.20235291

**Authors:** Laith Yakob

## Abstract

Self-instigated isolation is heavily relied on to curb SARS-CoV-2 transmission. Accounting for uncertainty in the latent and prepatent periods, as well as the proportion of infections that remain asymptomatic, the limits of this intervention at different phases of infection resurgence are estimated. We show that by October 2020, SARS-CoV-2 transmission rates in England had already begun exceeding levels that could be interrupted using this intervention alone, lending support to the second national lockdown on November 5^th^ 2020.

---

A general population lockdown occurred in England on 23^rd^ March 2020 to reduce SARS-CoV-2 transmission. This drastic intervention successfully inhibited disease spread by rapidly depleting the opportunities for transmission events between infected and susceptible people remaining in general circulation [1].

Subsequent to easing out of lockdown from July 4^th^ 2020, infections resurged and England entered its second national lockdown on November 5^th^ 2020. The return of millions of (largely susceptible) people to general circulation underlies the epidemic re-entering an exponential growth phase. However, also culpable in the current public health emergency is the failure of interventions during the period following lockdown’s release.

Contact tracing endeavours in 2020 to reduce SARS-CoV-2 transmission have been ineffective in England and so isolation has been primarily instigated by those responding to symptoms’ development in themselves or their close associations [2]. The mechanism by which this reactive isolation operates is importantly distinct from pre-emptive mass quarantine (lockdown). Symptoms-prompted, reactive isolation only applies to individuals who are infected (c.f. the total population), and, more specifically, to those who register symptoms. Hence infectious individuals who have not yet experienced symptoms, or who will never experience them, are missed.

The mathematical epidemiology of reactive isolation is fairly nascent yet critical in the context of the current epidemic. Here, we generate estimates for reactive isolation thresholds that account for uncertainties in the latent and pre-patent period of infection as well as in the proportion of infected individuals that register and respond appropriately to symptoms.

## Mathematical derivation of reactive self-isolation

Beginning with the simplest derivation for physical isolation: the pre-emptive quarantine threshold proportion (*Q*) is *Q* > (1 – (1/*R*)) where ‘*R*’ is the reproduction number [3]. For reactive isolation (*Q\**), this threshold is inflated to account for the leaked infections occurring because of the delay between becoming infectious and first exhibiting symptoms: *Q\** > (1 – (1/*R*)) x [1/(1 – ((*p* – *l*)/*g*))]. Respectively, *p* and *l* are the prepatent and latent period of infection (in days), and *g* is time until recovery (12 days on average [4]). If symptoms typically develop at the same time as an individual becomes infectious, the square-bracket component equals one and the original threshold (*Q*) is regained. A further modification can be made to account for the proportion of infections that never give rise to symptoms (denoted ‘*a*’): *Q\*\** > (1/(1 - *a*)) x (1 – (1/*R*)) x [1/(1 – ((*p* – *l*)/*g*))]. For example, if half of infections remained asymptomatic, the proportion of symptomatic infections that need to be isolated to achieve an equivalent impact must be doubled. As with those who never develop symptoms, individuals who fail to respond appropriately to developing symptoms – early indication is that this is not a negligible proportion [5] – will continue to contribute to transmission, so ‘*a*’ could be considered a composite of these two proportions.

## Accounting for uncertainty in parametrization

The latent and prepatent periods are quite variable for COVID-19 patients. Instead of single point estimates for these parameters, collated data form a distribution of reported times. The latent period is drawn at random from a Weibull distribution and then subtracted from the random draw from a second Weibull distribution depicting the range of reported prepatent periods. Fig 1A illustrates these distributions as informed by the clinical and epidemiological literature [6-8]. Also shown is the distribution of times between development of infectiousness and symptoms onset as fitted to 10,000 random draws. The distributions of prepatent and latent periods overlap so to avoid the possibility of symptoms developing prior to infectiousness, random draws whereby infectiousness trailed the day of symptoms onset were removed and resampled. 10,000 random draws were then made from this newly derived distribution of the delay between infectiousness and symptoms, and the isolation threshold (Q**) was estimated for a range of *R* values and a range of asymptomatic proportions (Python code for this analysis is freely available on https://github.com/lwyakob/COVIDquarantine).

**Figure 1.**
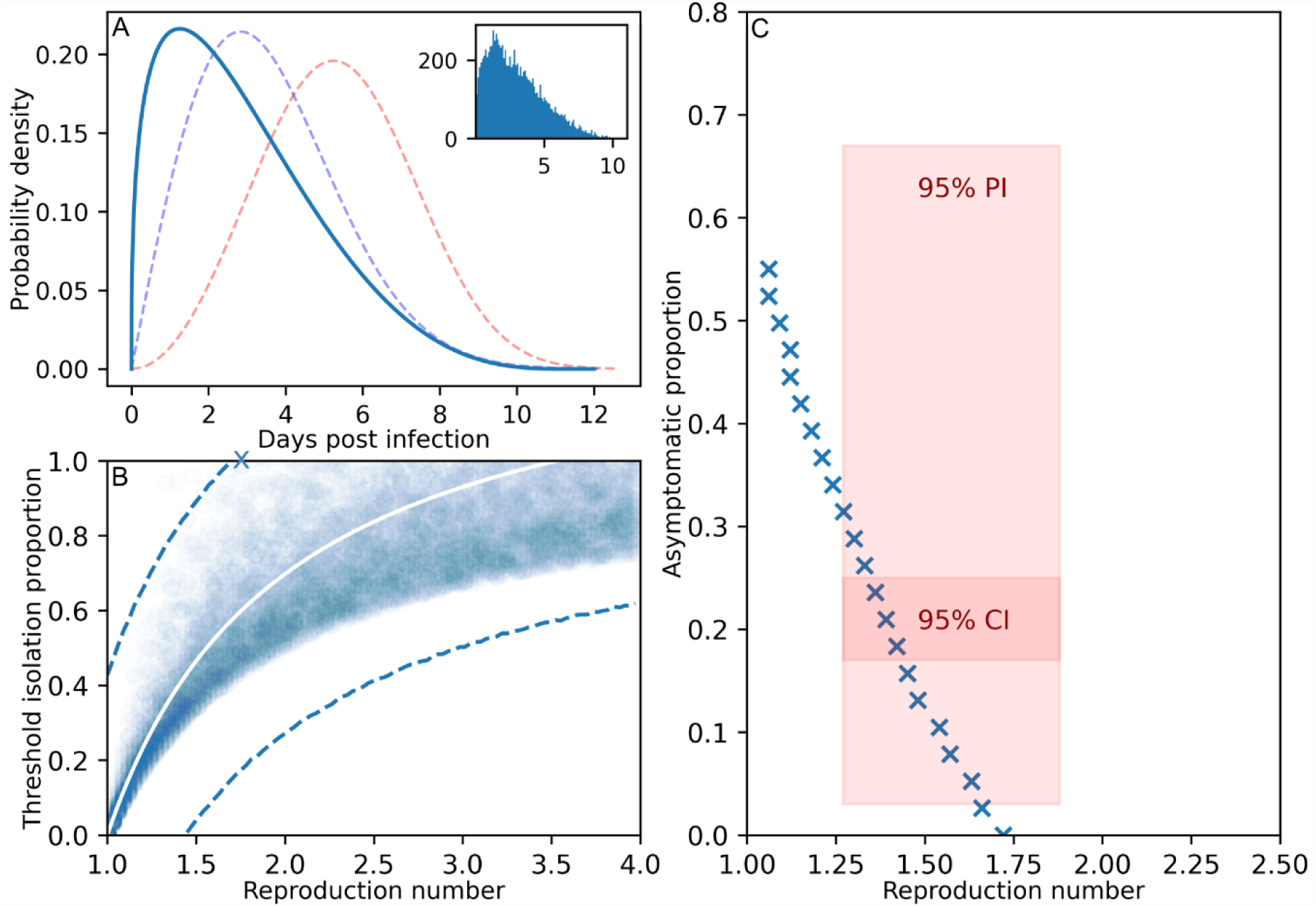
**A) Dashed lines indicate distributions for the latent (blue, Weibull(α=4, β=2)) and prepatent period (red, Weibull(α=6, β=3)) as derived from the COVID-19 literature [6-8]. The solid line is the resulting distribution for the time difference between the two from which 10,000 random draws were made (inset). B) The isolation threshold (*Q\**) as calculated for the 10,000 random draws along with the mean (white line) and 95% predictive interval (dashed lines). The blue cross indicates the theoretical maximum *R* number for which reactive isolation may interrupt transmission. C) The maximum asymptomatic proportion of COVID-19 infections that permits transmission interruption by reactive isolation for a range of *R* values (the hatched curve is calculated using the expression for Q**). The red boxes illustrate estimates for the asymptomatic proportion and the *R* for England as of October 2020 [9, 10].**

## Isolation thresholds accounting for uncertainty

Fig 1B shows the mean isolation threshold required to control SARS-CoV-2 accounting for the range of estimates for the prepatent and latent periods. The value for *R* is dynamic, varying according to current intervention effectiveness and population-level susceptibility, so the isolation threshold is shown for a range of plausible *R* values. The form of the relationship between *Q\** and *R* shows an isolation threshold that increases asymptotically with reproduction number. However, allowing for uncertainty in prepatent and latent periods results in a wide 95% prediction interval. The interpretation is that when accounting for both the uncertainty in estimating the population mean, plus the random variation of the individual values, reactive isolation cannot interrupt transmission (at least 95 times out of 100) if *R* already exceeds a value of ∼1.7 (blue cross on Fig 1B marks the *R* value whereby the isolation threshold proportion exceeds unity).

Reactive isolation is further limited when asymptomatic infections comprise a non-negligible proportion (alternatively, when those exhibiting symptoms fail to isolate themselves to some degree). Fig 1C shows the theoretical limits of the proportion of infections that can be asymptomatic and yet SARS-CoV-2 transmission interrupted through isolating symptomatic individuals (using the Q** expression). Superimposed on this trade-off between the reproduction number and the isolation threshold are estimates for *R* in England as of October 2020 [10], and the 95% confidence and predictive intervals for the proportion of infections that remain asymptomatic as generated by a living systematic review [9]. Respectively, by October 75% and 85% of these parameter spaces were already beyond the level at which reactive isolation can be sufficient to interrupt transmission (i.e., these regions fall to the right of the hatched arc in Fig 1C meaning the isolation threshold proportion exceeds unity).

## Limitations and future work

One limitation of the current analysis is the consideration of transmission and control at the population level rather than stratified by various risk factors. To address this, results were generated for a full range of *R* values. It is important to note that stratification would impact the derivation of *R* but not the population-level isolation thresholds calculated for a given *R* value [11]. Another limitation is the implicit assumption that, in the absence of intervention, asymptomatically infected individuals contribute to onwards transmission as much as symptomatically infected individuals. It is unclear how questionable this assumption is but clinical studies indicate that asymptomatic and symptomatic individuals have similar viral loads [12]. Should evidence arise of their differential contributions to transmission, the model and code associated with this study can be modified easily to account for this feature.

Even during pre-emptive quarantine (i.e., lockdown) the formulae described here continue to apply to those who remain in general circulation (e.g., essential personnel). Future work should look at how isolation thresholds can be estimated to inform this intervention combination, among others.

## Data Availability

Available from : https://github.com/lwyakob/COVIDquarantine
The link is in the manuscript.

## Data availability statement

Data and model code are all available from https://github.com/lwyakob/COVIDquarantine

